# Differential Impact of Mitigation Policies and Socioeconomic Status on COVID-19 Prevalence and Social Distancing in the United States

**DOI:** 10.1101/2020.10.20.20216119

**Authors:** Hsien-Yen Chang, Wenze Tang, Elham Hatef, Christopher Kitchen, Jonathan P. Weiner, Hadi Kharrazi

## Abstract

**Background:** The spread of COVID-19 has highlighted the long-standing health inequalities across the U.S. as neighborhoods with fewer resources were associated with higher rates of COVID-19 transmission. Although the stay-at-home order was one of the most effective methods to contain its spread, residents in lower-income neighborhoods faced barriers to practicing social distancing. We aimed to quantify the differential impact of stay-at-home policy on COVID-19 transmission and residents’ mobility across neighborhoods of different levels of socioeconomic disadvantage.

**Methods:** This was a comparative interrupted time-series analysis at the county level. We included 2,087 counties from 38 states which both implemented and lifted the state-wide stay-at-home order. Every county was assigned to one of four equally-sized groups based on its levels of disadvantage, represented by the Area Deprivation Index. Prevalence of COVID-19 was calculated by dividing the daily number of cumulative confirmed COVID-19 cases by the number of residents from the 2010 Census. We used the Social Distancing Index, derived from the COVID-19 Impact Analysis Platform, to measure the social distancing practice. For the evaluation of implementation, the observation started from Mar 1 ^St^ 2020 to one day before lifting; and, for lifting, it ranged from one day after implementation to Jul 5 ^th^ 2020. We calculated a comparative change of daily trends in COVID-19 prevalence and Social Distancing Index between counties with three highest disadvantage levels and those with the least level before and after the implementation and lifting of the stay-at-home order, separately.

**Results:** On both stay-at-home implementation and lifting dates, COVID-19 prevalence was much higher among counties with the highest or lowest disadvantage level, while mobility decreased as the disadvantage level increased. Mobility of the most disadvantaged counties was least impacted by stay-at-home implementation and relaxation compared to counties with the most resources; however, disadvantaged counties experienced the largest relative increase in COVID-19 infection after both stay-at-home implementation and relaxation.

**Conclusions:** Neighborhoods with varying levels of socioeconomic disadvantage reacted differently to the implementation and relaxation of COVID-19 mitigation policies. Policymakers should consider investing more resources in disadvantaged counties as the pandemic may not stop until most neighborhoods have it under control.

## Background

In the US, the total number of confirmed COVID-19 cases has skyrocketed from 30 patients on Mar 1 ^St^, 2020 to over 6.4mil on Sep 11 with total deaths exceeding 192k.[1] With no vaccines available for2020 to over 6.4mil on Sep 11 ^th^ with total deaths exceeding 192k.[1] With no vaccines available for prevention and no proven high-efficacy drug treatments, physicians can only provide supportive care for COVID-19 patients.[2] The lack of a definite treatment has also propelled healthcare professionals and policymakers to further focus their efforts on preventing the transmission of the disease.

Various approaches to slowing down the COVID-19 transmission have been recommended, with a focus on either self-protection or limited in-person contacts.[3, 4] In the absence of a universal mitigation policy by the US federal government, state and local governments have implemented a range of social distancing policies to restrict in-person contacts and limit mobility, such as restricting dine-in at restaurants, closing non-essential business, and banning large gatherings.[3-5] Among a wide range of mitigation policies, the stay-at-home (SAH) order has been the most restrictive policy with early studies documenting various levels of effectiveness of such policy.[5-9]

Forty states, including the District of Columbia, implemented the state-wide SAH order in Mar and Apr 2020 during an initial surge of COVID-19 transmission.[10] Despite the effectiveness of SAH order in reducing the COVID-19 transmission,[4-7, 9] its impact on economic activity was deemed detrimental.[4] Consequently, as COVID-19 transmission started to slow down between Apr and Jun 2020, 38 out of 40 states lifted their SAH order.[10] Evaluating the impact of the SAH orders, both implementation and relaxation of the orders, on COVID-19 transmission would provide useful information given the recent resurgence of COVID-19 transmissions across the US.[1]

The spread of COVID-19 has highlighted the established health inequalities across the US. For example, neighborhoods with higher income inequality, a higher proportion of racial or ethnic minorities, lower median family income, and higher unemployment rates were associated with higher rates of COVID-19 transmission, hospitalization, and death.[11-14] Similar associations were observed at the individual level.[15] Studies also found that residents in lower-income neighborhoods faced barriers to practicing physical distancing or following mobility restriction, particularly given the need to work outside the home.[16-22] These findings raise the question about the differential impact of the implementation and lifting of SAH orders on neighborhoods with varying levels of disadvantage and uneven levels of COVID-19 burden.

This study assesses the effectiveness of SAH policy on decreasing the COVID-19 transmission across neighborhoods with different levels of socioeconomic disadvantage represented by the Area Deprivation Index (ADI). We describe the differential impact of the implementation and lifting of SAH order on COVID-19 prevalence and residents’ mobility across counties with different levels of ADI (the highest/Q4, high/Q3, low/Q2, the lowest/Q1). To take into account the association of population density with social mobility and COVID-19 morbidity,[23-25] we perform additional analyses stratified by the population density.

## Methods

### Data

We compiled data from several data sources for this analysis. We derived the daily number of cumulative confirmed COVID-19 cases at the county level from the COVID-19 dashboard by the Center for Systems Science and Engineering (CSSE) at Johns Hopkins University.[4] We used the Social Distancing Index (SDI) to measure the social distancing practice at the county level. We derived the daily SDI from the COVID-19 Impact Analysis Platform (CIAP) created by the Maryland Transportation Institute at the University of Maryland.[26] We determined the SES of a county using ADI.[27, 28] We derived the county-specific characteristics, such as the percentage of the elderly, from the US Census American Community Survey (ACS) data of 2018. We derived the population size of each county from the 2010 Census. We did not obtain institutional review board approval due to the use of publicly available, de-identified data, per usual institutional policy.

### Social Distancing Index (SDI)

SDI uses the location data from anonymized mobile devices that are integrated with geographical population data. SDI for each state and county was derived from information such as percentage of people who are staying home, the average number of trips per person and average distance traveled by each person.[26] SDI, ranging from 0 to 100, represented the extent residents/visitors would practice social distancing: “0” indicated no social distancing in the community while “100” indicated all residents were staying at home and no visitors were entering the county. The mean national county-level SDI was 34.1 on March 7 ^th^ when SDI data first became available.

### Area Deprivation Index (ADI)

ADI is a widely used measure of socioeconomic disadvantage at various geographical levels, and contains factors related to employment, income, education, and housing condition.[27-29] We took the following steps to calculate the ADI using the latest Census data: (1) we used 71 independent census variables from 5-year estimate American Community Survey data of 2018 to construct 17 ADI grouped variables; (2) we summed up weighted 17 ADI components as total scores using Singh et al. methodology;[29] and, (3) we normalized total scores to have a mean of 100 and a standard deviation of 20. We constructed the ADI raw scores for 52 states (i.e., continental states, Alaska, Hawaii, and Puerto Rico) at the county level and assigned each county to one of four equally-sized ADI levels based on its ADI score (the highest/Q4: 76 ^th^ −100 ^th^ percentile of the ADI score, high/Q3: 51 ^st^ to 75 ^th^ percentile, low/Q2: 26 ^th^ to 50 ^th^ percentile, and the lowest/Q1 ^st^: 1 to 25 ^th^ percentile).

### State Stay-At-Home Order

Out of 51 states including the District of Columbia, 11 states did not implement SAH order (Arkansas, Connecticut, Iowa, Kentucky, Nebraska, North Carolina, Oklahoma, South Dakota, Texas, Utah, and Wyoming) and 13 states did not lift SAH order as of Aug 3 ^rd^ 2020 (the aforementioned 11 states plus California and New Mexico).[10] To keep the inference population consistent between analyses of SAH implementation and lifting effects, we restricted all of our analyses to the 38 states with both implementing and lifting SAH orders, covering 2,087 counties and approximately 212 out of 338 (62.5%) million Americans. The highest ADI score (i.e., Q4) indicated the most disadvantaged neighborhoods, while lower ADI scores (Q3 to Q1) indicated lower levels of disadvantages accordingly.

## Outcomes

We included two outcomes in the analysis: COVID-19 prevalence and SDI. For each county, we calculated the daily COVID-19 prevalence by dividing the daily number of confirmed COVID-19 cases by the number of residents. We derived the daily SDI at the county level directly from the CIAP. To minimize the fluctuations in the derived SDI, we used 7-day simple-averaged SDI (Figure 2).

### Time Segments

We used two different observation periods for the comparative analysis: (1) To assess the effect of the SAH implementation, we included the observation period starting from March 1 ^st^ to one day before the SAH lifting date; and, (2) To assess the effect of the SAH lifting, we included the observation period starting from one day after the SAH implementation date to Jul 5 ^th^ (data cutoff date) of each county. We obtained the dates of implementation and lifting of SAH order at the state level from the “COVID-19 US state policy database” project by Raifman et al. at the Boston University School of Public Health (reviewed on Aug 3 ^rd^, 2020).[10] We assigned the counties within the same state to the same dates of SAH implementation and lifting.

### Analyses

We first described and compared the county characteristics within each of the four ADI levels using chi-squared tests for categorical variables and Kruskal-Wallis test for continuous variables. To visualize the change in trends of county-level COVID-19 prevalence and SDI, we plotted these two outcomes by calendar time, averaging over counties of the same ADI level. We produced separate plots for period around implementation and lifting of SAH order due to potential scale differences in outcomes. We also added time frame indicating the earliest and last SAH implementation/lifting to each plot.

To compare the policy impact across counties at four ADI levels, we adopted a comparative interrupted time series (ITS) framework. The comparative ITS analysis has been used to evaluate the policy impact on outcomes between the case (with the policy implementation) and the control (without the policy implementation).[30, 31] In our study, these observations provided a comparative change of daily trends in the outcomes between counties at three higher ADI levels (Q2-Q4) and those at the lowest ADI level (Q1; the reference group) before and after the implementation/lifting of SAH order. In order to take into account either the anticipated or delayed effect of SAH order, we chose all actual interrupted time points after examining plots of empirical trends of COVID-19 prevalence and SDI at SAH implementation and lifting index date (Appendix Figure 1a and 1b).[32] Thus, we used the following dates to evaluate the respective policy’ impact on COVID-19 prevalence: 5 days after SAH implementation and 20 days after SAH lifting date. Similarly, we used 15 days before SAH implementation and 40 days before SAH lifting to evaluate their respective impact on change in SDI.

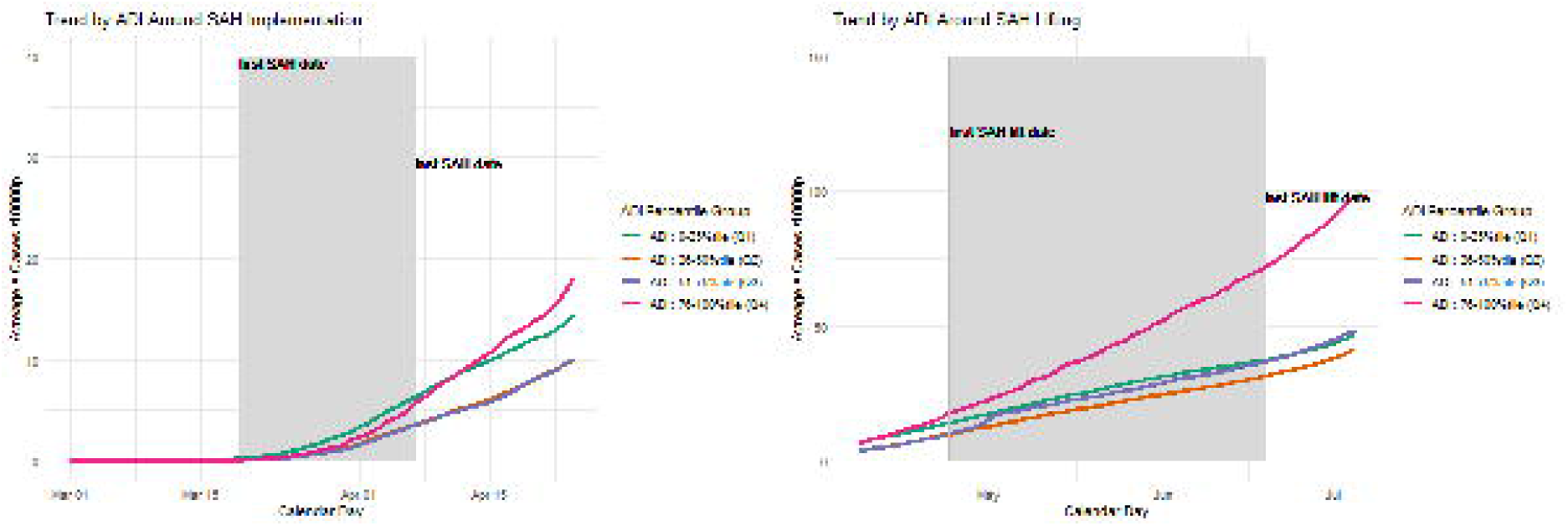

We used a linear mixed-effect model to quantify the effect of implementing and lifting SAH order on each outcome. Our parameter of interest was the interaction of the ADI level indicator and the post-policy day indicator, representing the effect of the policies on the rate of change (trend) from the pre-policy to post-policy periods across four ADI levels. For each county, we included random effect terms for intercept (e.g., baseline COVID-19 prevalence), pre-policy outcome trend (e.g., COVID-19 prevalence trend before SAH implementation) and post-policy outcome trend (e.g., difference in COVID-19 prevalence trend after SAH implementation). We also included day of week to adjust for its time-varying effect. All statistical tests were two-sided with alpha level of 0.05.

All analyses were conducted using R 4.0.

## Results

### Characteristics of counties by ADI level

We included 2,087 counties in the analysis, with 521 or 522 counties in each level of ADI. The observation time for SAH implementation ranged from 86.2 days among counties with ADI Q1 level to 95.8 days among counties with ADI Q3 level. The observation time for SAH lifting was about the same, ranging from 89.5 days among counties with ADI Q1 level to 93.2 days among counties with ADI Q4 level. We observed statistically significant differences in characteristics of counties across four levels of ADI (Table 1). As the county’s ADI level decreased, the population size, the population density, the median family income and the percentage of residents with at least high school diploma increased while the percentages of families in poverty and residents unemployed decreased. For example, the mean number of residents increased from 37,381 in ADI Q4 level to 176,972 in ADI Q1 level. The mean age ranged from 40.7 years old among counties with ADI Q4 level to 42.3 years old among counties with ADI Q2 level.

**Table 1:**
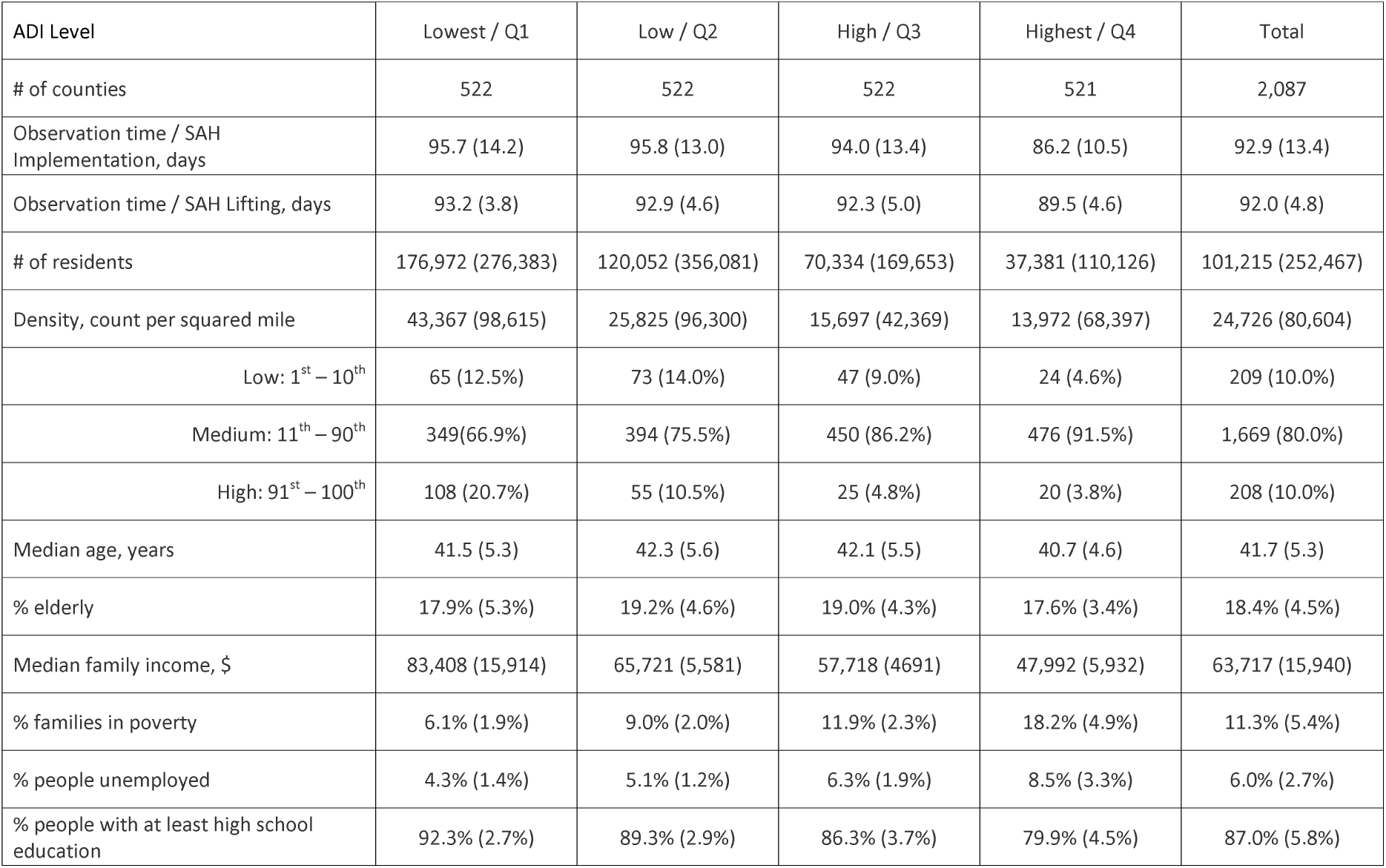
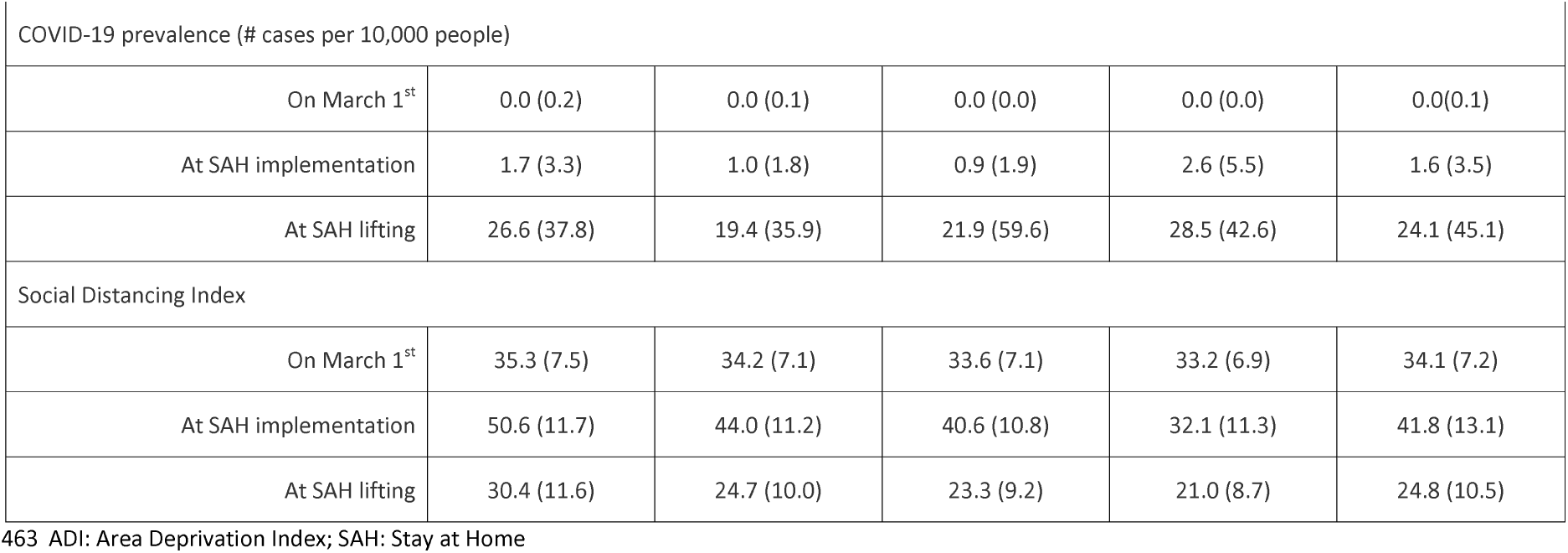
Characteristics of counties by the Area Deprivation Index level

| ADI Level | Lowest / Q1 | Low / Q2 | High / Q3 | Highest / Q4 | Total |
| --- | --- | --- | --- | --- | --- |
| # of counties | 522 | 522 | 522 | 521 | 2,087 |
| Observation time / SAH Implementation, days | 95.7 (14.2) | 95.8 (13.0) | 94.0 (13.4) | 86.2 (10.5) | 92.9 (13.4) |
| Observation time / SAH Lifting, days | 93.2 (3.8) | 92.9 (4.6) | 92.3 (5.0) | 89.5 (4.6) | 92.0 (4.8) |
| # of residents | 176,972 (276,383) | 120,052 (356,081) | 70,334 (169,653) | 37,381 (110,126) | 101,215 (252,467) |
| Density, count per squared mile | 43,367 (98,615) | 25,825 (96,300) | 15,697 (42,369) | 13,972 (68,397) | 24,726 (80,604) |
| Low: 1<sup>st</sup> – 10<sup>th</sup> | 65 (12.5%) | 73 (14.0%) | 47 (9.0%) | 24 (4.6%) | 209 (10.0%) |
| Medium: 11<sup>th</sup> – 90<sup>th</sup> | 349(66.9%) | 394 (75.5%) | 450 (86.2%) | 476 (91.5%) | 1,669 (80.0%) |
| High: 91<sup>st</sup> – 100<sup>th</sup> | 108 (20.7%) | 55 (10.5%) | 25 (4.8%) | 20 (3.8%) | 208 (10.0%) |
| Median age, years | 41.5 (5.3) | 42.3 (5.6) | 42.1 (5.5) | 40.7 (4.6) | 41.7 (5.3) |
| % elderly | 17.9% (5.3%) | 19.2% (4.6%) | 19.0% (4.3%) | 17.6% (3.4%) | 18.4% (4.5%) |
| Median family income, \$ | 83,408 (15,914) | 65,721 (5,581) | 57,718 (4691) | 47,992 (5,932) | 63,717 (15,940) |
| % families in poverty | 6.1% (1.9%) | 9.0% (2.0%) | 11.9% (2.3%) | 18.2% (4.9%) | 11.3% (5.4%) |
| % people unemployed | 4.3% (1.4%) | 5.1% (1.2%) | 6.3% (1.9%) | 8.5% (3.3%) | 6.0% (2.7%) |
| % people with at least high school education | 92.3% (2.7%) | 89.3% (2.9%) | 86.3% (3.7%) | 79.9% (4.5%) | 87.0% (5.8%) |
| COVID-19 prevalence (# cases per 10,000 people) |  |  |  |  |  |
| On March 1<sup>st</sup> | 0.0 (0.2) | 0.0 (0.1) | 0.0 (0.0) | 0.0 (0.0) | 0.0(0.1) |
| At SAH implementation | 1.7 (3.3) | 1.0 (1.8) | 0.9 (1.9) | 2.6 (5.5) | 1.6 (3.5) |
| At SAH lifting | 26.6 (37.8) | 19.4 (35.9) | 21.9 (59.6) | 28.5 (42.6) | 24.1 (45.1) |
| Social Distancing Index |  |  |  |  |  |
| On March 1<sup>st</sup> | 35.3 (7.5) | 34.2 (7.1) | 33.6 (7.1) | 33.2 (6.9) | 34.1 (7.2) |
| At SAH implementation | 50.6 (11.7) | 44.0 (11.2) | 40.6 (10.8) | 32.1 (11.3) | 41.8 (13.1) |
| At SAH lifting | 30.4 (11.6) | 24.7 (10.0) | 23.3 (9.2) | 21.0 (8.7) | 24.8 (10.5) |

463 ADI: Area Deprivation Index; SAH: Stay at Home

A U-shape relationship was observed between ADI level and the COVID-19 prevalence (Table 1). Counties with ADI Q4 or Q1 level had much higher COVID-19 prevalence on both SAH implementation and lifting dates than those with Q3 or Q2 level. A dose-response relationship was identified between the ADI level and SDI: as the ADI level increased (i.e., more disadvantaged neighborhoods), SDI decreased on both SAH order implementation and lifting dates.

### Average county-level COVID-19 prevalence by the ADI level over calendar time

Figure 1 depicts the average county-level COVID-19 prevalence by the ADI level over calendar time. At the SAH implementation, counties with ADI Q4 level had the largest increase in the slope of COVID-19 prevalence from the pre-to post-SAH implementation, while counties with ADI Q1 level seemed to have the smallest increase. Eventually, counties with ADI Q4 level had the highest absolute COVID-19 prevalence rate. Counties with ADI Q3 or Q2 level had similar prevalence rates across the entire observation period. Similarly, at the SAH lifting, counties with ADI Q4 level also had the largest increase in the slope of COVID-19 prevalence from the pre-to post-SAH lifting. Counties with other three ADI levels had similar trends before and after SHA lifting.

### Average county-level SDI by the ADI level over calendar time

Figure 2 shows the average county-level SDI by the ADI level over calendar time. SDI showed more fluctuations than COVID-19 prevalence. SDI across all counties increased before, stayed flat during, and started to decrease after the SAH implementation. SDI across all counties decreased before, continued to decrease during, and started to increase after the SAH lifting. In counties with higher ADI levels, SDI was lower over the entire observation period.

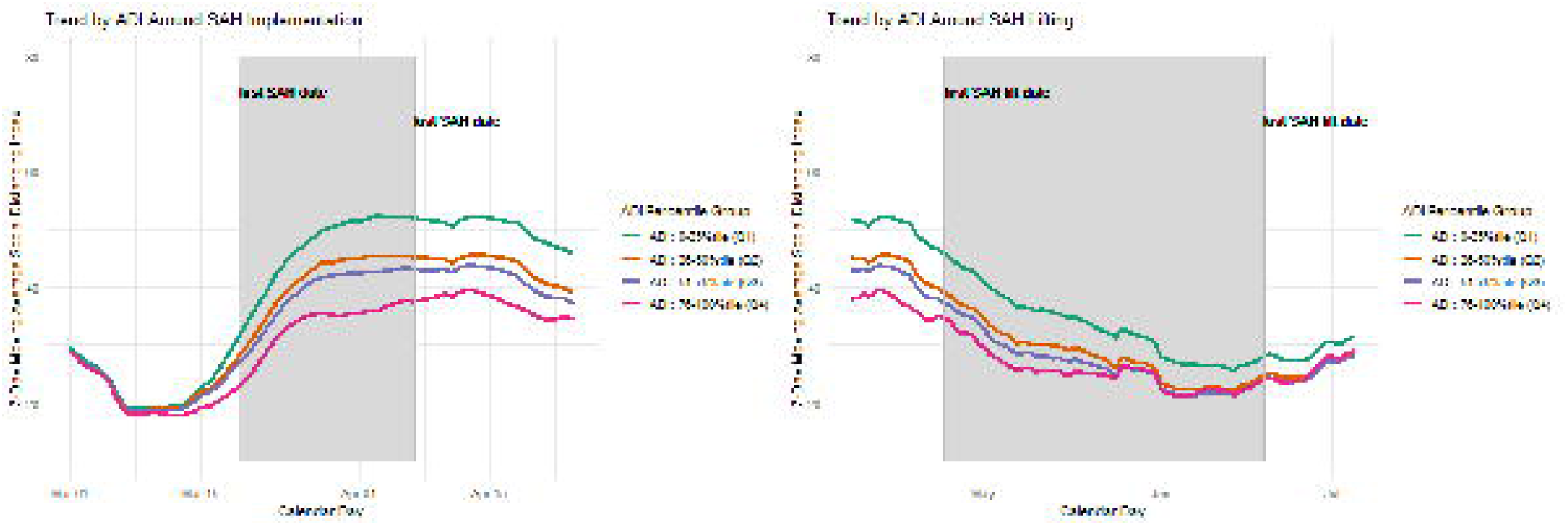

### Effect of SAH Implementation

Following the implementation of the SAH order, the counties with ADI Q4 level experienced a statistically significantly relative increase in the daily trend of COVID-19 prevalence (0.371 prevalence/day, 95% Confidence Interval (CI) 0.211 to 0.532) compared to those with ADI Q1 level. The counties with ADI Q2 or Q3 level did not experience such significantly relative differences (Figure 3 & Appendix Table 1).

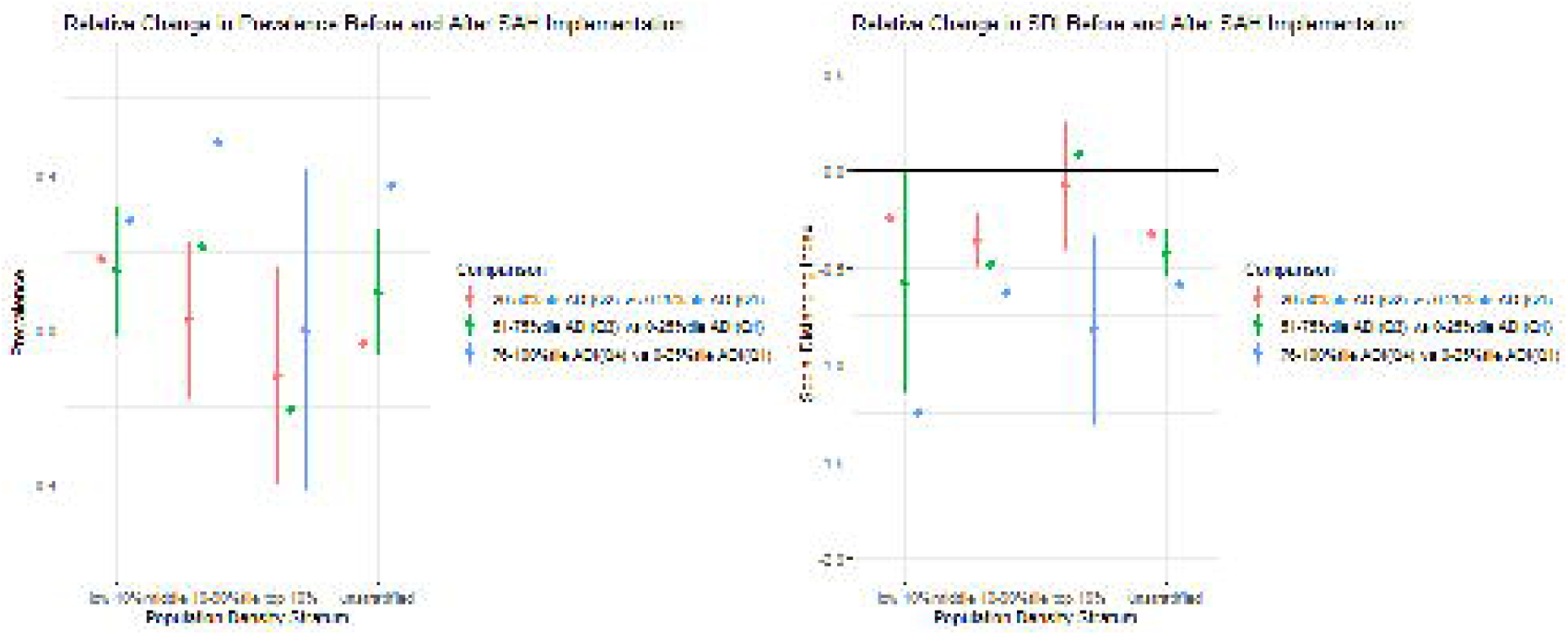

Counties with the non-Q1 ADI levels experienced statistically significantly relative reductions in the daily trend of SDI compared to those with ADI Q1 level. For example, a daily relative decline of 0.592 SDI/day (95% CI −0.717 to −0.467) were detected when comparing the counties with ADI Q4 to ADI Q1 level. Compared to the counties with ADI Q1 level, such relative reduction was 0.335 SDI/day (95% CI −0.454 to −0.215) for those with ADI Q2 level and 0.429 SDI/day (95% CI −0.549 to −0.308) for those with ADI Q3 level (Figure 3 & Appendix Table 1).

Results from the stratified analyses by the population density were mostly similar to those from the unstratified analysis of the entire study population with some exceptions. For example, in the analyses of the daily prevalence, no statistically significantly associations were observed among high-density counties while the significant association was also observed comparing ADI Q2 to ADI Q1 level (0.181 prevalence/day, 95% CI 0.031 to 0.331) among low-density counties and ADI Q3 to ADI Q1 level (0.212 prevalence/day, 95% CI 0.016 to 0.408) among medium-density counties. In the analyses of daily SDI, no significant associations were observed comparing ADI Q2 to ADI Q1 level among low-density counties and comparing ADI Q2 or Q3 to ADI Q1 level among high-density counties (Figure 3 & Appendix Table 1).

### Effect of SAH Lifting

Following the lifting of the SAH order, the counties with ADI Q4 level experienced a statistically significantly relative increase in the daily trend of COVID-19 prevalence (0.449 prevalence/day, 95% CI 0.280 to 0.618) compared to those with ADI Q1 level. Counties with ADI Q2 or Q3 level did not experience such significantly relative differences (Figure 4 & Appendix Table 2).

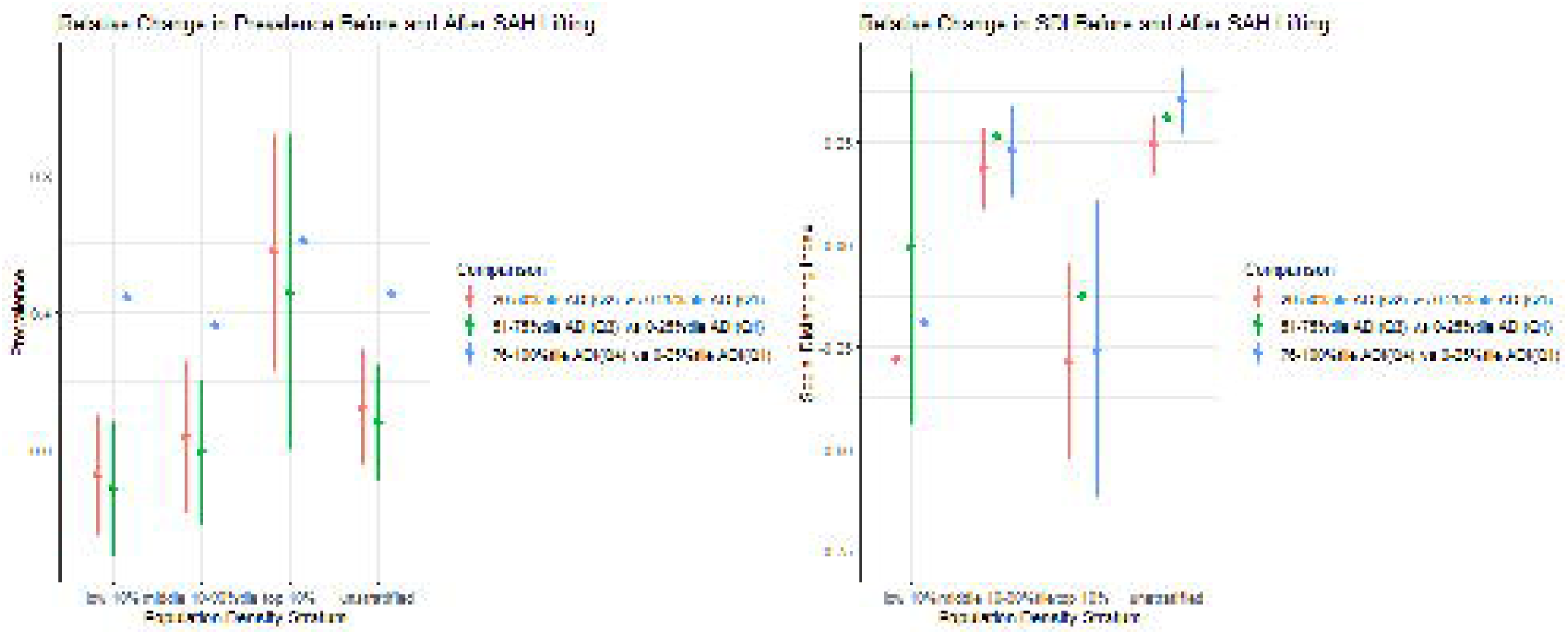

Counties with ADI non-Q1 levels (i.e., Q2, Q3 and Q4) experienced statistically significantly relative increases in the daily trend of SDI compared to those with ADI Q1 level. For example, there was a daily relative increase of 0.352 SDI/day (95% CI 0.272 to 0.433) comparing the counties with ADI Q4 to Q1 level. Compared to the counties with ADI Q1 level, such relative increase was 0.243 SDI/day (95% CI 0.171 to 0.315) for those with ADI Q2 level and 0.310 SDI/day (95% CI 0.237 to 0.383) for those with ADI Q3 level (Figure 4 & Appendix Table 2).

Results from the density-stratified analyses were similar to those from the unstratified analysis of the entire study population for the outcome of COVID-19 prevalence but not SDI. For example, no statistically significant relative difference was detected in daily SDI comparing low-density counties with the three ADI non-Q1 levels to those with Q1 level. The only statistically significant relative difference among high-density counties was the daily relative reduction of 0.287 SDI/day (95% CI −0.529 to −0.045) comparing ADI Q2 to Q1 level (Figure 4 & Appendix Table 2).

## Discussion

COVID-19 has affected US neighborhoods and communities disparately, with minorities and less resourceful communities taking most of the burden. Despite these initial findings, evidence on the effect of neighborhood-level disparities on the effectiveness of SAH policy and COVID-19 transmission is still lacking. Our study revealed the role of social disparities in compliance with the SAH order implementation and relaxation and the impact on COVID-19 transmission across neighborhoods. When compared to the counties with ADI Q1 level, we found a comparative increase in the COVID-19 rate among counties with ADI Q4 level and relatively less increase in the social distancing among counties with higher ADI after the SAH order was implemented. After the lifting of the SAH order, compared to the counties with ADI Q1 level, we found a comparative increase in the COVID-19 rate among counties with ADI Q4 level and relatively more social distancing among counties with higher ADI. In short, mobility of counties with ADI Q4 level (i.e., most disadvantaged counties) was least impacted by SAH implementation and lifting compared with Q1 level, but experienced the worst relative increase in COVID-19 infection after both SAH implementation and lifting.

The differential responses to the implementation of the SAH order by neighborhoods’ levels of socioeconomic disadvantage have been reported in previous studies, but most studies focused on the mobility rate among residence of different neighborhoods.[16, 17, 20, 21] One study found that physical distancing orders were associated with less increase in staying home in low-income vs. high-income neighborhoods (1.5% vs 2.4%).[17] Another study found that areas with fewer resources had more subway use in New York City.[20] These findings were consistent with ours. It is likely that residents in more disadvantaged neighborhoods (represented by higher ADI levels) had fewer resources to stay at home. For example, they may still need to work outside the home such as essential workers. Consequently, such comparative increase in residents’ mobility may lead to an increase in the in-person contacts, which eventually may have resulted in the relative increase in the COVID-19 prevalence in disadvantaged neighborhoods (i.e., high ADI), as we observed in this study. Moreover, social distancing might be more challenging in socioeconomically disadvantaged neighborhoods with high housing density and overcrowding.[33-35] Additionally, communities that are mainly comprised of economically challenged households are more likely to be exposed to COVID-19 due to their overrepresentation in the low-wage, essential work at the front lines.[33]

We have not identified any research on the impact of lifting the SAH orders. The SAH order lifting might have increased social and entertainment activities, with residents in the low ADI neighborhoods (i.e., less disadvantaged) having the means to socialize compared to those in high ADI neighborhoods. On the other hand, more people in the high ADI neighborhoods (i.e., higher disadvantaged) may have lost their jobs during the pandemic (decrease in mobility given no need to go out to work) or have to continue working regardless of the SAH order, such as essential workers (no change in mobility). Thus, we observed relatively increases in social distancing after such lifting among higher disadvantaged neighborhoods. However, even with the relatively decreasing trend in mobility, neighborhoods with the highest ADI still experienced a relatively increasing rate of COVID-19 cases. It is possible that residents in less disadvantaged neighborhoods practiced self-protection measures better (e.g., able to purchase face masks or hand sanitizers), as various studies have shown that higher-resource neighborhoods were associated with less COVID-19 transmission.[11-14] Our findings also align with another study that pointed out health disparities may play a more important role in COVID-19 transmission than government interventions (such as SAH order) and community-level compliance to such interventions.[13]

Our study had some limitations. First, this was an ecological study and results from the aggregated data might not be generalizable to individuals. Second, some counties implemented various policies aiming at reducing COVID-19 transmission during our study period (e.g., shelter-in-place or stay-at-home orders; restricting dine-in at restaurants; closing nonessential business such as bars, entertainment venues, and gyms; banning large social gatherings; and closing public schools). Although we adopted the random intercept method to control for the fixed state effects, our results may not be attributed to the SAH order alone. Third, we used ADI to represent the disadvantage level of a neighborhood (representing the overall social determinants of health), and SDI to represent mobility. While both measures, especially ADI, have been examined extensively, these measures may still not capture the concepts they aim to represent completely. For example, SDI is based on mobile device data; it may not capture the mobility in the extremely disadvantaged counties with low use of smartphones and/or low penetration of internet.

## Conclusions

Our study, despite having limitations, showed that neighborhoods with varying levels of disadvantage reacted differently to the implementation and relaxation of COVID-19 mitigation policies. Those with the highest ADI (i.e., most disadvantaged counties) observed the largest relative increases after both the SAH implementation and lifting. Policymakers should consider investing more resources in these disadvantaged counties to help them contain the COVID-19 transmission in future SAH implementations and liftings, as the pandemic may not stop until most, if not all, neighborhoods have it under control.

## Data Availability

The data file is uploaded as an appendix.

## List of abbreviations

ACS: American Community Survey
ADI: Area Deprivation Index
ITS: Interrupted time series
CIAP: COVID-19 Impact Analysis Platform
CSSE: Center for Systems Science and Engineering
SAH: Stay-at-home
SDI: Social Distancing Index

## DECLARATIONS

### Ethics approval and consent to participate

The datasets we used are publicly available and de-identified information at the country level; therefore, ethics review board approval for this study is not required.

### Consent for publication

All authors have approved this manuscript for publication.

### Availability of data and material

All data generated or analyzed during this study are included as a supplementary compressed csv file named “df_SAH_policy_on_covid19.zip”.

### Authors’ contributions

HC and WT designed the study, managed data, did analyses, and drafted the manuscript. CK obtained data, provided critical comments and revised the manuscript. EH, JP and HK provided critical comments and revised the manuscript.

## Acknowledgements

Not applicable.

## Reference

1. COVID-19 Map [https://coronavirus.jhu.edu/map.html]

2. Coronavirus COVID-19 (SARS-CoV-2) Trusted Information at the Point of Care Guides [https://www.hopkinsguides.com/hopkins/view/Johns_Hopkins_ABX_Guide/540747/all/Coronavirus_COVID_19__SARS_CoV_2_]

3. Coronnavirus Disease 2019 (COVID-19) - Your Health [https://www.cdc.gov/coronavirus/2019-ncov/your-health/index.html]

4. Juneau C-E, Pueyo T, Bell M, Gee G, Collazzo P, Potvin L: Evidence-Based, Cost-Effective Interventions To Suppress The COVID-19 Pandemic: A Systematic Review. medRxiv 2020:2020.2004.2020.20054726.

5. Teslya A, Pham TM, Godijk NG, Kretzschmar ME, Bootsma MCJ, Rozhnova G: Impact of self-imposed prevention measures and short-term government-imposed social distancing on mitigating and delaying a COVID-19 epidemic: A modelling study. medRxiv 2020:2020.2003.2012.20034827.

6. Dreher N, Spiera Z, McAuley FM, Kuohn L, Durbin JR, Marayati NF, Ali M, Li AY, Hannah TC, Gometz A et al: Impact of policy interventions and social distancing on SARS-CoV-2 transmission in the United States. medRxiv 2020:2020.2005.2001.20088179.

7. Fowler JH, Hill SJ, Obradovich N, Levin R: The Effect of Stay-at-Home Orders on COVID-19 Cases and Fatalities in the United States. medRxiv 2020:2020.2004.2013.20063628.

8. Furukawa NW, Brooks JT, Sobel J: Evidence Supporting Transmission of Severe Acute Respiratory Syndrome Coronavirus 2 While Presymptomatic or Asymptomatic. Emerg Infect Dis 2020, 26(7).

9. Koh WC, Naing L, Wong J: Estimating the impact of physical distancing measures in containing COVID-19: an empirical analysis. medRxiv 2020:2020.2006.2011.20128074.

10. COVID-19 US state policy database [www.tinyurl.com/statepolicies]

11. Abedi V, Olulana O, Avula V, Chaudhary D, Khan A, Shahjouei S, Li J, Zand R: Racial, Economic and Health Inequality and COVID-19 Infection in the United States. medRxiv 2020:2020.2004.2026.20079756.

12. Bhowmik T, Tirtha SD, Iraganaboina NC, Eluru N: A Comprehensive Analysis of COVID-19 Transmission and Fatality Rates at the County level in the United States considering Socio- Demographics, Health Indicators, Mobility Trends and Health Care Infrastructure Attributes. medRxiv 2020:2020.2008.2003.20164137.

13. Jalali AM, Khoury SG, See J, Gulsvig AM, Peterson BM, Gunasekera RS, Buzi G, Wilson J, Galbadage T: Delayed Interventions, Low Compliance, and Health Disparities Amplified the Early Spread of COVID-19. medRxiv 2020:2020.2007.2031.20165654.

14. Robertson LS: COVID-19 Confirmed Cases and Fatalities in 883 U.S. Counties with a Population of 50,000 or More: Predictions Based on Social, Economic, Demographic Factors and Shutdown Days. medRxiv 2020:2020.2006.2025.20139956.

15. Rozenfeld Y, Beam J, Maier H, Haggerson W, Boudreau K, Carlson J, Medows R: A model of disparities: risk factors associated with COVID-19 infection. Int J Equity Health 2020, 19(1):126.

16. Dasgupta N, Jonsson Funk M, Lazard A, White BE, Marshall SW: Quantifying the social distancing privilege gap: a longitudinal study of smartphone movement. medRxiv 2020:2020.2005.2003.20084624.

17. Jay J, Bor J, Nsoesie E, Lipson SK, Jones DK, Galea S, Raifman J: Neighborhood income and physical distancing during the COVID-19 pandemic in the U.S. medRxiv 2020:2020.2006.2025.20139915.

18. Shadmi E, Chen Y, Dourado I, Faran-Perach I, Furler J, Hangoma P, Hanvoravongchai P, Obando C, Petrosyan V, Rao KD et al: Health equity and COVID-19: global perspectives. International Journal for Equity in Health 2020, 19(1):104.

19. Silva DS, Smith MJ: Social distancing, social justice, and risk during the COVID-19 pandemic. Canadian Journal of Public Health 2020, 111(4):459–461.

20. Sy KTL, Martinez ME, Rader B, White LF: Socioeconomic disparities in subway use and COVID- 19 outcomes in New York City. medRxiv 2020.

21. Torres TS, Hoagland B, Bezerra DRB, Garner A, Jalil EM, Coelho LE, Benedetti M, Pimenta C, Grinsztejn B, Veloso VG: Impact of COVID-19 Pandemic on Sexual Minority Populations in Brazil: An Analysis of Social/Racial Disparities in Maintaining Social Distancing and a Description of Sexual Behavior. AIDS Behav 2020.

22. Hatef E, Chang H-Y, Kitchen C, Weiner JP, Kharrazi H: Assessing the Impact of Neighborhood Socioeconomic Characteristics on COVID-19 Prevalence Across Seven States in the United States. Frontiers in Public Health 2020, 8:354.

23. Krishnamachari B, Dsida A, Zastrow D, Harper B, Morris A, Santella A: Effects of Government Mandated Social Distancing Measures on Cumulative Incidence of COVID-19 in the United States and its Most Populated Cities. medRxiv 2020:2020.2005.2022.20110460.

24. Machida M, Nakamura I, Saito R, Nakaya T, Hanibuchi T, Takamiya T, Odagiri Y, Fukushima N, Kikuchi H, Amagasa S et al: The actual implementation status of self-isolation among Japanese workers during the COVID-19 outbreak. Trop Med Health 2020, 48:63.

25. You H, Wu X, Guo X: Distribution of COVID-19 Morbidity Rate in Association with Social and Economic Factors in Wuhan, China: Implications for Urban Development. Int J Environ Res Public Health 2020, 17(10).

26. Zhang L, Ghader S, Pack ML, Xiong C, Darzi A, Yang M, Sun Q, Kabiri A, Hu S: AN INTERACTIVE COVID-19 MOBILITY IMPACT AND SOCIAL DISTANCING ANALYSIS PLATFORM. medRxiv 2020:2020.2004.2029.20085472.

27. Kind AJH, Buckingham WR: Making Neighborhood-Disadvantage Metrics Accessible - The Neighborhood Atlas. N Engl J Med 2018, 378(26):2456–2458.

28. 2015 Area Deprivation Index v2.0 [https://www.neighborhoodatlas.medicine.wisc.edu/]

29. Singh GK: Area deprivation and widening inequalities in US mortality, 1969-1998. Am J Public Health 2003, 93(7):1137–1143.

30. Rutkow L, Chang HY, Daubresse M, Webster DW, Stuart EA, Alexander GC: Effect of Florida’s Prescription Drug Monitoring Program and Pill Mill Laws on Opioid Prescribing and Use. JAMA Intern Med 2015, 175(10):1642–1649.

31. Chang HY, Lyapustina T, Rutkow L, Daubresse M, Richey M, Faul M, Stuart EA, Alexander GC: Impact of prescription drug monitoring programs and pill mill laws on high-risk opioid prescribers: A comparative interrupted time series analysis. Drug Alcohol Depend 2016, 165:1–8.

32. Bernal JL, Cummins S, Gasparrini A: Interrupted time series regression for the evaluation of public health interventions: a tutorial. Int J Epidemiol 2017, 46(1):348–355.

33. Bibbins-Domingo K: This Time Must Be Different: Disparities During the COVID-19 Pandemic. Ann Intern Med 2020, 173(3):233–234.

34. Bowleg L: We’re Not All in This Together: On COVID-19, Intersectionality, and Structural Inequality. Am J Public Health 2020, 110(7):917.

35. Labor Force Statistics from the Current Population Survey [https://www.bls.gov/cps/cpsaat11.htm]

