## Appendix Table for "Differential Impact of Mitigation Policies and Socioeconomic Status on COVID-19 Prevalence and Social Distancing in the United States"

Table 1: Effect of SAH implementation by ADI level and population density

| Outcome | Population Density Stratum | ADI level (ref: Q1) | Relative difference in trend |
| --- | --- | --- | --- |
| COVID-19 Prevalence | Unstratified | Q2 | -0.035  (-0.195, 0.126) |
|  |  | Q3 | 0.097  (-0.063, 0.258) |
|  |  | Q4 | 0.371  (0.211, 0.532) |
|  | Low | Q2 | 0.181  (0.031, 0.331) |
|  |  | Q3 | 0.151  (-0.017, 0.319) |
|  |  | Q4 | 0.284  (0.074, 0.494) |
|  | Medium | Q2 | 0.026  (-0.176, 0.228) |
|  |  | Q3 | 0.212  (0.016, 0.408) |
|  |  | Q4 | 0.485  (0.291, 0.679) |
|  | High | Q2 | -0.120  (-0.401, 0.162) |
|  |  | Q3 | -0.208  (-0.586, 0.169) |
|  |  | Q4 | -0.002  (-0.415, 0.412) |
| Social Distancing Index | Unstratified | Q2 | -0.335  (-0.454, -0.215) |
|  |  | Q3 | -0.429  (-0.549, -0.308) |
|  |  | Q4 | -0.592  (-0.717, -0.467) |
|  | Low | Q2 | -0.248  (-0.760, 0.263) |
|  |  | Q3 | -0.585  (-1.154, -0.016) |
|  |  | Q4 | -1.248  (-1.993, -0.503) |
|  | Medium | Q2 | -0.361  (-0.496, -0.226) |
|  |  | Q3 | -0.487  (-0.619, -0.355) |
|  |  | Q4 | -0.634  (-0.768, -0.499) |
|  | High | Q2 | -0.080  (-0.414, 0.253) |
|  |  | Q3 | 0.082  (-0.368, 0.531) |
|  |  | Q4 | -0.823  (-1.305, -0.342) |

ADI: Area Deprivation Index; SAH: Stay at Home

Table 2: Effect of SAH lifting by ADI level and population density

| Outcome | Population Density Stratum | ADI level (ref: Q1) | Relative difference in trend |
| --- | --- | --- | --- |
| COVID-19 Prevalence | Unstratified | Q2 | 0.122  (-0.049, 0.292) |
|  |  | Q3 | 0.079  (-0.092, 0.249) |
|  |  | Q4 | 0.449  (0.280, 0.618) |
|  | Low | Q2 | -0.075  (-0.247, 0.097) |
|  |  | Q3 | -0.112  (-0.310, 0.086) |
|  |  | Q4 | 0.442  (0.201, 0.682) |
|  | Medium | Q2 | 0.038  (-0.180, 0.257) |
|  |  | Q3 | -0.007  (-0.218, 0.205) |
|  |  | Q4 | 0.361  (0.153, 0.569) |
|  | High | Q2 | 0.573  (0.230, 0.917) |
|  |  | Q3 | 0.454  (-0.003, 0.911) |
|  |  | Q4 | 0.605  (0.099, 1.110) |
| Social Distancing Index | Unstratified | Q2 | 0.243  (0.171, 0.315) |
|  |  | Q3 | 0.310  (0.237, 0.383) |
|  |  | Q4 | 0.352  (0.272, 0.433) |
|  | Low | Q2 | -0.281  (-0.669, 0.106) |
|  |  | Q3 | -0.008  (-0.441, 0.424) |
|  |  | Q4 | -0.193  (-0.764, 0.377) |
|  | Medium | Q2 | 0.186  (0.085, 0.287) |
|  |  | Q3 | 0.264  (0.163, 0.364) |
|  |  | Q4 | 0.228  (0.118, 0.338) |
|  | High | Q2 | -0.287  (-0.529, -0.045) |
|  |  | Q3 | -0.126  (-0.438, 0.187) |
|  |  | Q4 | -0.260  (-0.622, 0.103) |

ADI: Area Deprivation Index; SAH: Stay at Home
