## Supplementary figures and images for "Differential Impact of Mitigation Policies and Socioeconomic Status on COVID-19 Prevalence and Social Distancing in the United States"

### 1a. Average Prevelance by ADI Groups followup day.png

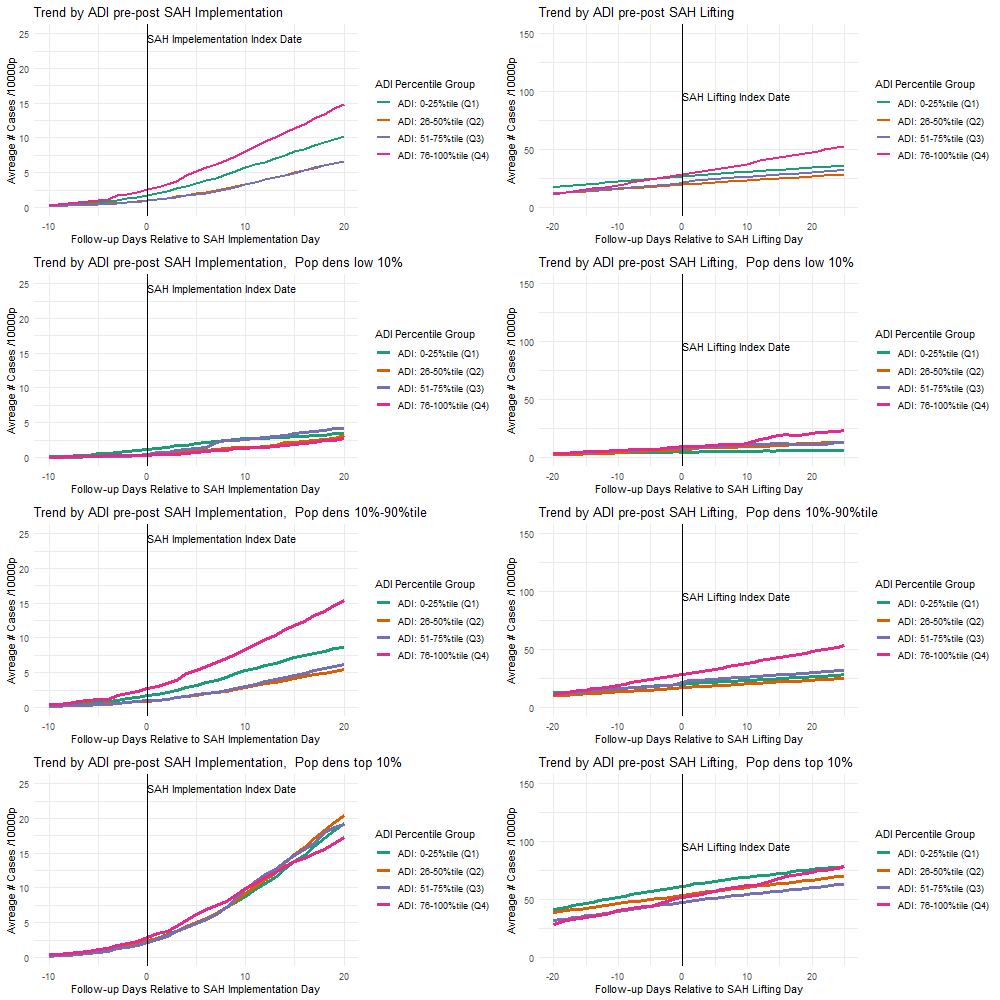

### 1b. 7-Day Moving Average SDI by ADI Groups followup day.png

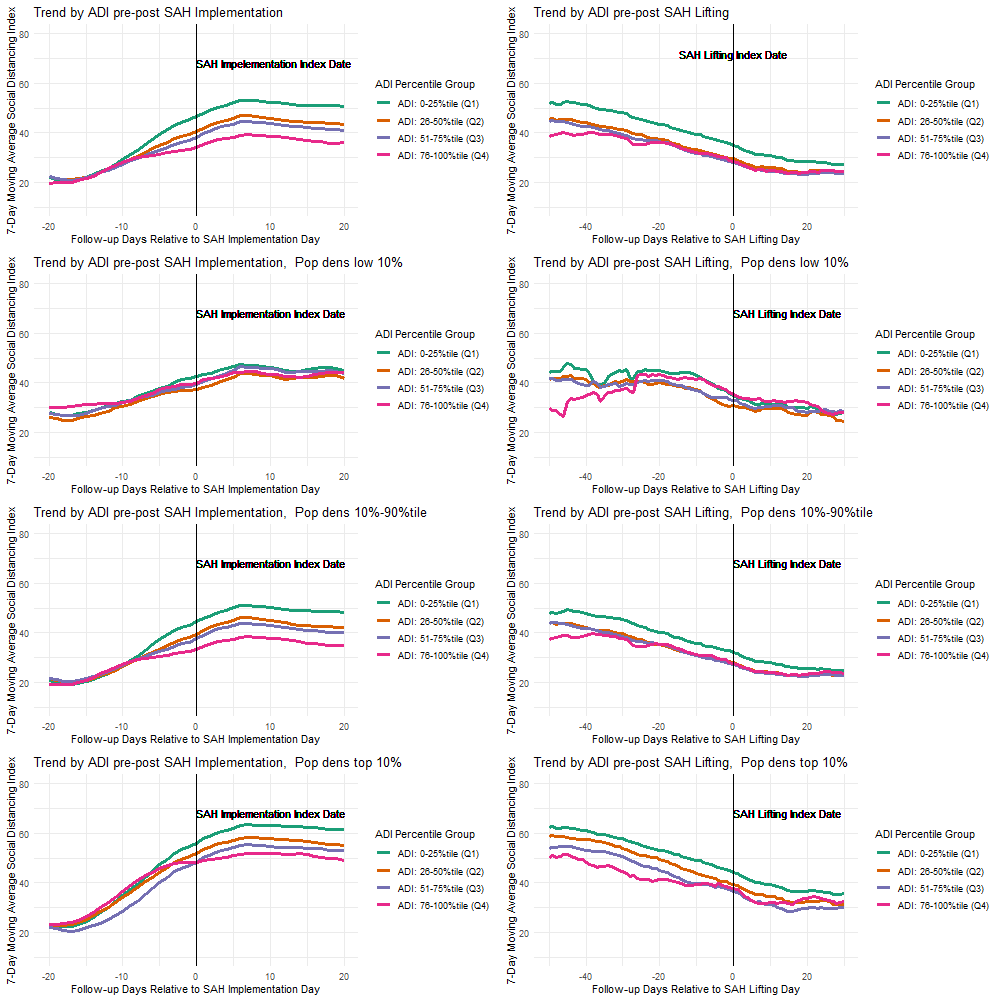

### 2a. Average Prevelance by ADI Groups calendar day.png

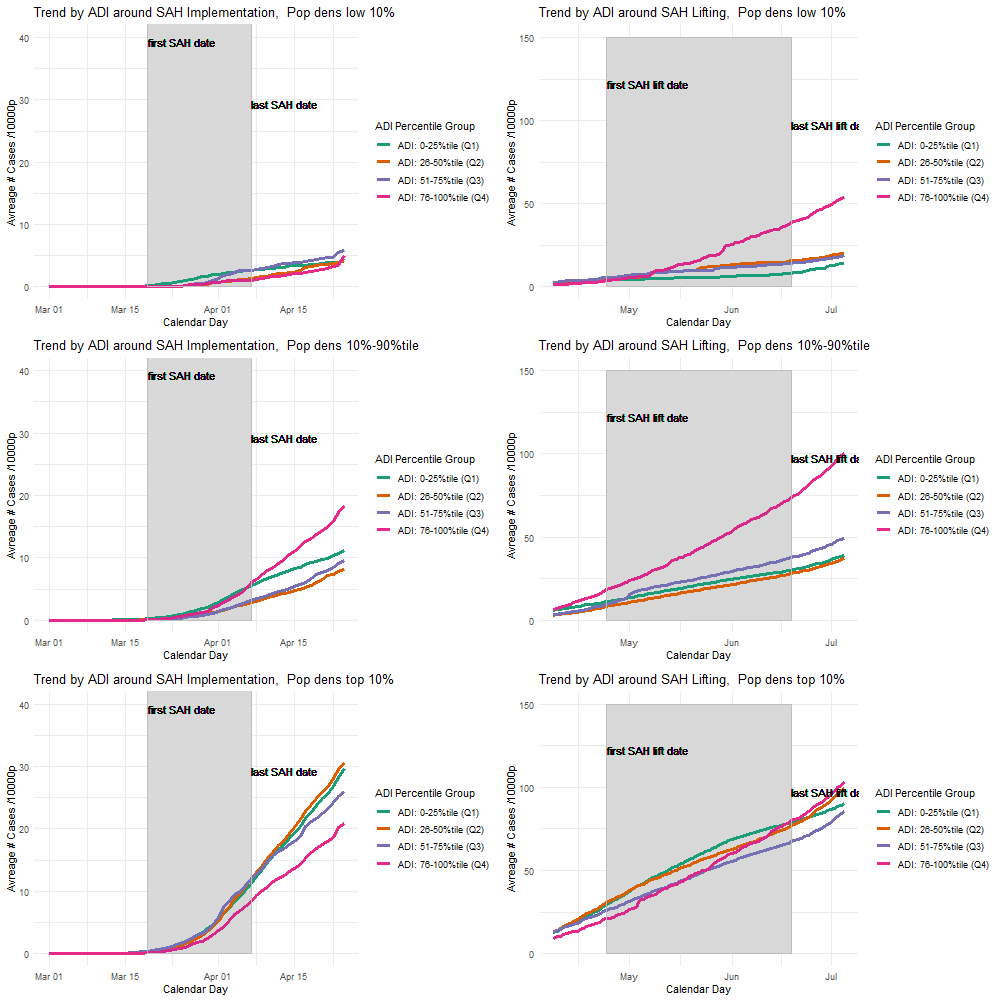

### 2b. 7-Day Moving Average SDI by ADI Groups calendar day.png

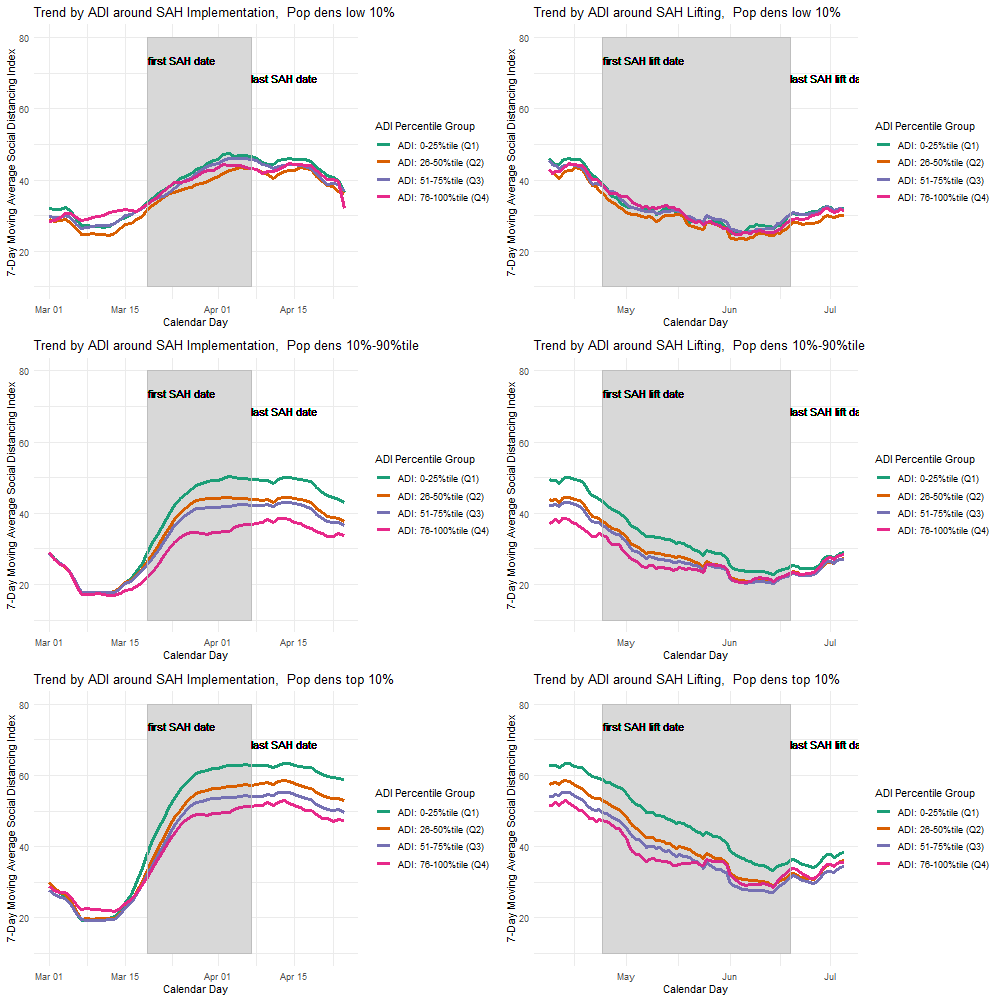
